# Emergence and spread of a sub-lineage of SARS-CoV-2 Alpha variant B.1.1.7 in Europe, and with further evolution of spike mutation accumulations shared with the Beta and Gamma variants

**DOI:** 10.1101/2021.11.01.21265749

**Authors:** Marlena Stadtmüller, Alexa Laubner, Fabian Rost, Sylke Winkler, Eva Patrasová, Lenka Šimůnková, Susanne Reinhardt, Johanna Beil, Alexander H. Dalpke, Buqing Yi

## Abstract

SARS-CoV-2 evolution plays a significant role in shaping the dynamics of the COVD-19 pandemic. To monitor the evolution of SARS-CoV-2 variants, **t**hrough international collaborations, we performed genomic epidemiology analyses on a weekly basis with SARS-CoV-2 samples collected from a border region between Germany, Poland and the Czech Republic in a global background. For identified virus mutant variants, active viruses were isolated and functional evaluations were performed to test their replication fitness and neutralization sensitivity against vaccine elicited serum neutralizing antibodies. Thereby we identified a new B.1.1.7 sub-lineage carrying additional mutations of nucleoprotein G204P and open-reading-frame-8 K68stop. Of note, this B.1.1.7 sub-lineage is the predominant B.1.1.7 variant in several European countries, such as Czech Republic, Austria and Slovakia. The earliest samples belonging to this sub-lineage were detected in November 2020 in a few countries in the European continent, but not in the UK. We have also detected its further evolution with extra spike mutations D138Y and A701V, which are signature mutations shared with the Beta and Gamma variants, respectively. Antibody neutralization assay of virus variant isolations has revealed that the variant with extra spike mutations is 3.2-fold less sensitive to vaccine-elicited antibodies as compared to other B.1.1.7 variants tested, indicating potential for immune evasion, but it also exhibited reduced replication fitness. The wide spread of this B.1.1.7 sub-lineage was related to the pandemic waves in early 2021 in various European countries. These findings about the emergence, spread, evolution, infection and transmission abilities of this B.1.1.7 sub-lineage add to our understanding about the pandemic development in Europe, and could possibly help to prevent similar scenarios in future.

## Introduction

As one of the SARS-CoV-2 variants of concern (VOC) (1), the alpha variant B.1.1.7 was first detected in the UK in September 2020. This variant was shown to be more transmissible (2-4) compared to previously detected other variants. In Europe, B.1.1.7 accounted for the majority of COVID-19 cases from February to May in spring 2021. The originally reported B.1.1.7 was characterized by 17 mutations including amino acid replacements and deletions on the spike protein (S), open-reading-frame-1ab (ORF1ab), open-reading-frame-8 (ORF8) and the nucleoprotein (5). These mutations might play a role in ACE2 receptor binding or neutralizing antibody escape (6, 7). One recent study has investigated the spatial invasion dynamics of B.1.1.7 in the UK, and the results of this study indicated that early B.1.1.7 growth rates were related with lineage export frequencies from a dominant source location (8). It remains unclear how B.1.1.7 could quickly spread to all the countries in Europe in spite of the strict lockdown including restrictions on international travels since December 2020 in almost all the European countries.

In Germany, regular large-scale genomic surveillance was initiated in early January 2021. With SARS-CoV-2 samples sequenced locally and with virus genomes shared on GISAID (9), we have routinely carried out genomic epidemiology analyses to investigate local, national and international virus spreading conditions, and monitor the evolution of existing variants and the emergence of new variants. Through joint efforts in genomic surveillance with Czech and Poland partners, we detected that in the B.1.1.7 samples in the Czech Republic, around 95% samples carried two extra mutations: N_G204P and ORF8_68stop. In Germany, more than 30% of B.1.1.7 samples showed these two extra mutations as well. More detailed analysis of international samples revealed that this B.1.1.7-N:G204P-ORF8:K68stop sub-lineage has been widely distributed in Europe. Here, we describe the detection, characterization, transmission, evolution and functional analyses of this B.1.1.7 sub-lineage, and the spreading pattern of B.1.1.7 in Europe in January 2021 when B.1.1.7 got detected in most European countries. The relevant information may help to understand why and how the B.1.1.7 waves could take place across Europe in the spring of 2021, thereby possibly promoting suitable strategies for preventing the spread of other variants of concern that evolve quickly.

## Methods

### Establishment of genome sequence data set for emerging variant monitoring

We combined SARS-CoV-2 sequences generated from samples collected in a border region between Germany, Poland and Czech Republic, with full-length SARS-CoV-2 sequences periodically downloaded from GISAID (9) to build up genome sequence data set for emerging variant monitoring (locally generated sequences were shared on GISAID as well). We first performed quality check and filtered out low-quality sequences that met any of the following criteria: 1) sequences with less than 90% genome coverage; 2) genomes with too many mutations (defined as having >20 nucleotide mutations relative to the Wuhan reference), which would violate the SARS-CoV-2 molecular clock at the time of study; 3) genomes with more than ten ambiguous bases; and 4) genomes with clustered mutations, defined as mutations in close proximity to one another. These are the standard quality assessment parameters utilized in NextClade (https://clades.nextstrain.org). The current study was based on the 2.17 million global viral genomes available as of 30 June 2021.

### Lineage classification

We used the dynamic lineage classification method in this study through the Phylogenetic Assignment of Named Global Outbreak Lineages (PANGOLIN) software suite (https://github.com/hCoV-2019/pangolin) (10). This is intended for identifying the most epidemiologically important lineages of SARS-CoV-2 at the time of analysis (11).

### Phylogenetic analysis of SARS-CoV-2

Phylogenetic analysis was carried out to infer the transmission routes of B.1.1.7 in Europe (12) with a custom build of the SARS-CoV-2 NextStrain build (https://github.com/nextstrain/ncov) (13). The pipeline includes several Python scripts that manage the analysis workflow. Briefly, it allows for the filtering of genomes, the alignment of genomes in MAFFT (14), phylogenetic tree inference in IQ-Tree (15), tree dating (16) and ancestral state construction and annotation. The phylogeny analysis is rooted by Wuhan-Hu-1/2019.

### Epidemiology data

We analyzed daily cases of SARS-CoV-2 in the Czech Republic from publicly released data provided by the Ministry of Health of the Czech Republic (https://onemocneni-aktualne.mzcr.cz/covid-19), and 7-day incidence rates per 100K inhabitants were calculated accordingly based on the local population. Daily cases of SARS-CoV-2 in Poland was obtained from publicly released data provided by the Service of the Republic of Poland (https://www.gov.pl/web/koronawirus/wykaz-zarazen-koronawirusem-sars-cov-2), and 7-day incidence rates per 100K inhabitants were calculated accordingly as well.

### Viruses

All viruses used were patient isolates cultured from nasopharyngeal swabs. Virus stocks were grown on Vero E6 cells in DMEM GlutaMAX supplemented with 10% FBS, 1% non-essential amino acids and 1% penicillin/streptomycin. The second passage of each virus isolate was used for experiments. The virus isolates hCoV-19/Germany/SN-RKI-I-178035/2021 (similar to originally defined B.1.1.7, labelled as B.1.1.7_O), hCoV-19/Germany/SN-RKI-I-038776/2021 (B.1.1.7-N:G204P-ORF8:68stop sub-lineage, labelled as B.1.1.7_S), hCoV-19/Germany/SN-UKDD-91348010S+/2021(B.1.1.7-N:G204P-ORF8:68stop sub-lineage with extra spike mutations D138Y and A701V, labelled as B.1.1.7_S+) were used in the virus neutralization assay and growth kinetics measurement.

### Virus Neutralization Assay

All sera were derived from healthy individuals fully vaccinated with BNT162b2. A 2-fold dilution series of each serum was prepared in PBS+ (supplemented with 0.3 % bovine albumin, 1 mM MgCl_2_ and 1 mM CaCl_2_) and each serum concentration was incubated with 50 PFU of B.1.1.7_O, B.1.1.7_S or B.1.1.7_S+ for 1 h at 37°C. Confluent Vero E6 cells seeded the day before were infected with the virus-containing serum dilutions for 1 h at 37°C and 5% CO_2_ with occasional shaking. The inoculum was aspirated, cells washed with PBS and subsequently overlayed with semi-viscous Avicel Overlay Medium (double-strength DMEM, Avicel RC-581 in H2O 0.75 %, 10 % FCS, 0.01 % DEAE-Dextran and 0.05 % NaHCO_3_). After 3 days, cells were stained with 0.1 % crystal violet in 10 % formaldehyde and plaques were counted. ID_50_ values were calculated using GraphPad Prism 9.

### Virus Growth Kinetics

Calu 3 cells were seeded 3 days prior to infection. On the day of infection, cells were infected with B.1.1.7_O, B.1.1.7_S or B.1.1.7_S+ at MOI 0.1 diluted in PBS+ for 1h at 37°C and 5% CO_2_ with occasional shaking. Afterwards, the inoculum was aspirated, the cells were washed with PBS and fresh medium (DMEM GlutaMAX supplemented with 10% FBS, 1 % non-essential amino acids, 1% sodium pyruvate and 1% penicillin/streptomycin) was added. Supernatants were removed at 8, 16, 24, 48, 72 and 96 hours post infection (hpi). Infectious virus particles in the supernatant were determined using plaque assay, which was performed analogously to the neutralization assay from the infection step onwards. Results are given as plaque forming units (PFU) per ml. Graphs were generated using GraphPad Prism 9.

## Results

### 1. Identification of one specific B.1.1.7 sub-lineage with extra mutations in Europe

We evaluated 948,077 SARS-CoV-2 alpha variant genomes available as of June 30, 2021. One specific B.1.1.7 sub-lineage with two extra mutations: nucleoprotein G204P (**N_G204P)** and ORF8 K68stop (**Orf8_K68stop)** (Fig. 1A), was detected in various European countries. Surprisingly, this B.1.1.7-N:G204P-ORF8:68stop sub-lineage had very unequal distribution in Europe. In the Czech Republic, Hungary, Slovakia and Austria, this sub-lineage was predominant, accounting for around 60-95% of local B.1.1.7 related COVID-19 cases. In Germany, Denmark, Switzerland, Poland and France, this sub-lineage accounted for around 10-35% of B.1.1.7 related COVID-19 cases in each country (Fig. 1B). In Belgium and Netherlands, this variant only accounted for less than 10% of B.1.1.7 related cases. Of note, this B.1.1.7-N:G204P-ORF8:68stop sub-lineage comprised only 0.4% of total B.1.1.7 genomes in the UK.

**Fig. 1.**
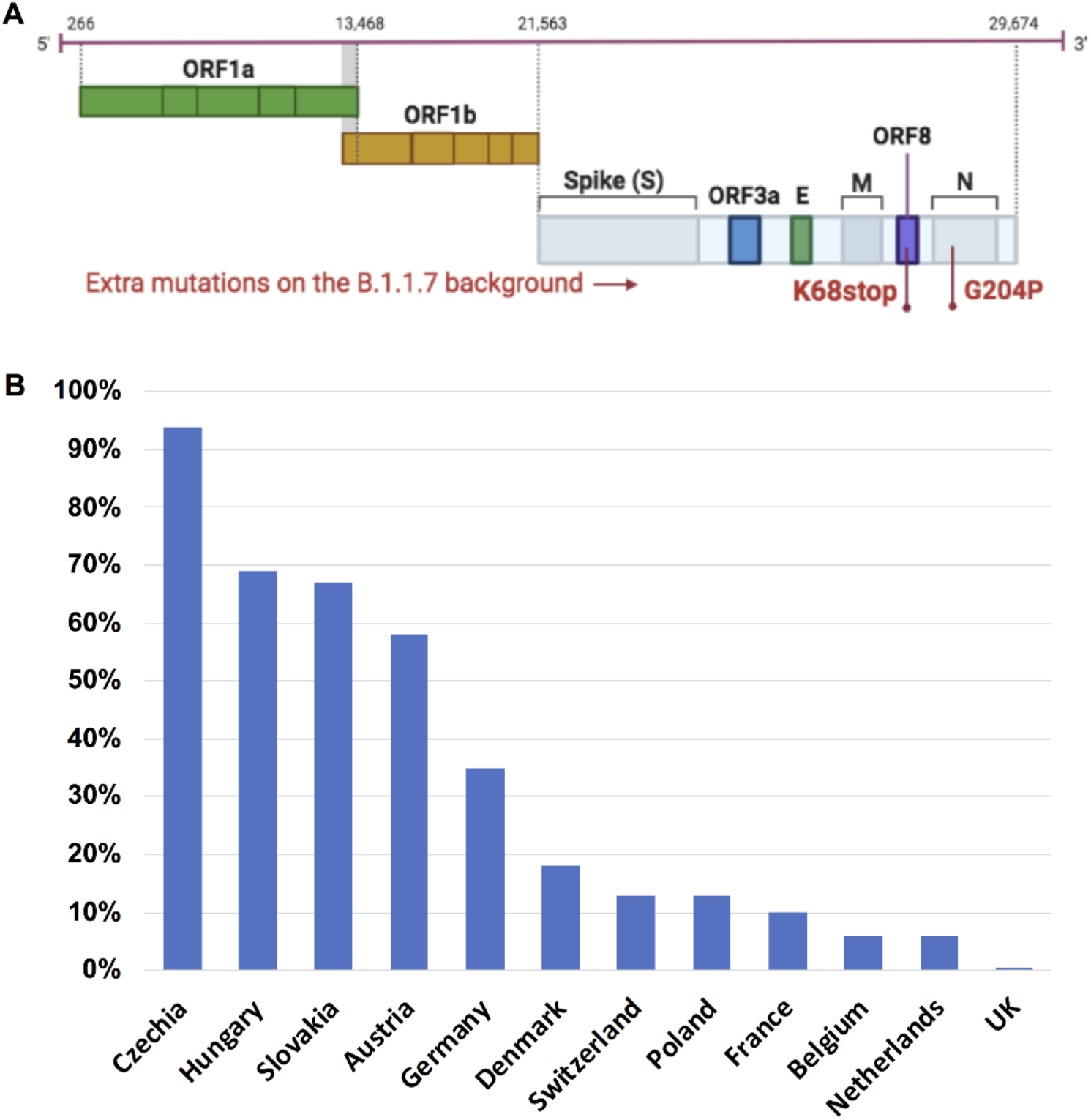
One specific B.1.1.7 sub-lineage with extra mutations was detected in Europe. **A**. This sub-lineage was characterized by two extra mutations: N_G204P and ORF8_K68stop. **B**. The distribution of this B.1.1.7-N:G204P-ORF8:68stop sub-lineage in Europe is shown as percentage of this sub-lineage in total B.1.1.7 in each country (data collected on June 30, 2021 including all available B.1.1.7 sequences on GISAID).

### 2. Estimated transmission routes of B.1.1.7 in Europe in January 2021

Since the spreading of B.1.1.7 was one critical driving force for the February-May wave in most European countries, we performed phylogeny analysis of B.1.1.7 to estimate the early transmission routes of B.1.1.7 in Europe. In most European countries, in January 2021 the B.1.1.7 already got frequently detected (17-19), so this analysis focused on the cross-country transmission taking place in January. Phylogeny analysis was performed with B.1.1.7 samples collected in January that are available at GISAID from 10 European countries (Austria: 202; Czech Republic: 161; UK: 57,847; Germany: 699; Switzerland: 1,117; Slovakia: 77; Italy: 373; Poland: 51; France: 1524; Denmark: 1412). In a few countries, especially in the UK, the sampling density was much higher than other countries, so we downsized the sample numbers to 100 randomly selected samples collected in January from each country. This condition was chosen because similar sample size from each country could largely prevent statistical errors in transmission route estimation. Fig. 2A shows the phylogeny-inferred cross-country transmission routes, which revealed two centers in the transmission network: UK and Czech Republic, indicating the frequencies of export from these two countries were much higher than that of other countries. Analysis of the transmission routes of the cluster of the B.1.1.7-N:G204P-ORF8:68stop sub-lineage (Fig. 2B) revealed that the Czech Republic was the center for this transmission network.

**Fig. 2.**
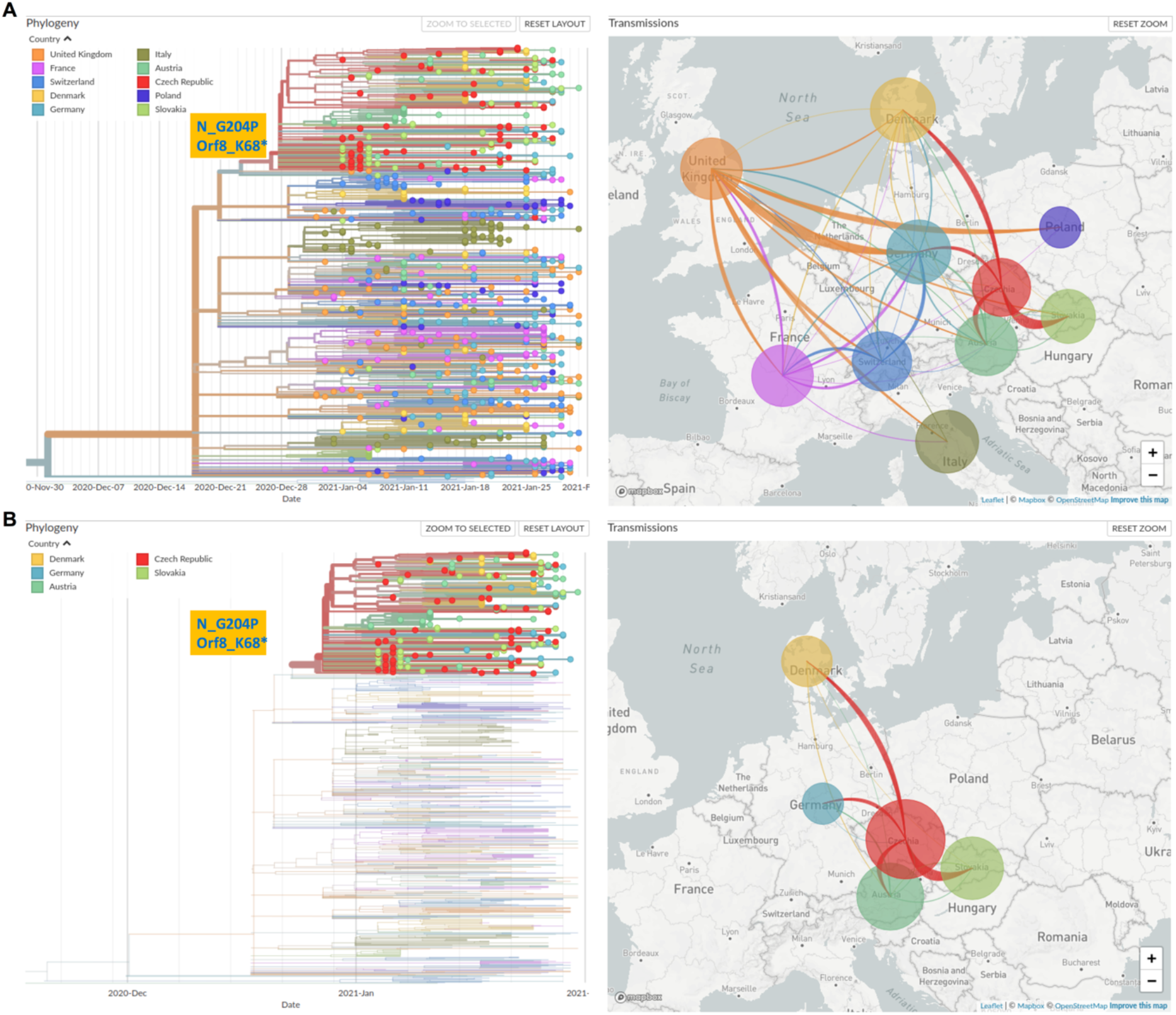
Transmission routes of B.1.1.7 in Europe in January 2021 inferred based on phylogeny analysis. The size of the circle represents the number of genomes from all B.1.1.7 (A) or from B.1.1.7-N:G204P-ORF8:68stop sub-lineage (B) in each country. The line colors correspond to the exporting locations. **A**. Left: Time and phylogeny tree of B.1.1.7 collected in January; Right: Estimated transmission routes of B.1.1.7 in Europe in January 2021. **B**. Left: Only the cluster characterized by N_G204P and ORF8_K68stop is displayed; Right: Estimated transmission routes of B.1.1.7 samples with the two extra mutations N_G204P and ORF8_K68stop (B.1.1.7-N:G204P-ORF8:68stop sub-lineage).

### 3. Exploration of the source of the B.1.1.7-N:G204P-ORF8:68stop sub-lineage

As this B.1.1.7 sub-lineage accounted for only 0.4% of B.1.1.7 related cases in UK till June 30, 2021 (Fig. 1), this finding suggests the UK was possibly not the direct source of this B.1.1.7 sub-lineage. To investigate the source and possible transmission routes of this B.1.1.7 sub-lineage at early stage, we analyzed all the B.1.1.7 samples belonging to this sub-lineage collected from all around Europe till end of January 2021. Earliest B.1.1.7 samples with N_G204P and Orf8_K68* occurred in November 2020 in the following countries: Switzerland, Austria, France, Slovakia and Denmark, with only one genome from each country being reported (Fig. 3A). Yet, till end of January 2021 it had been detected in 23 countries in Europe (Fig. 3B). Owing to the limited number of SARS-CoV-2 genomes that were sequenced last year, it is difficult to analyze more details about the early transmission routes of this sub-lineage. This B.1.1.7-N:G204P-ORF8:68stop sub-lineage was almost exclusive to Europe, and only a few samples were detected in Asia and North America.

**Fig. 3.**
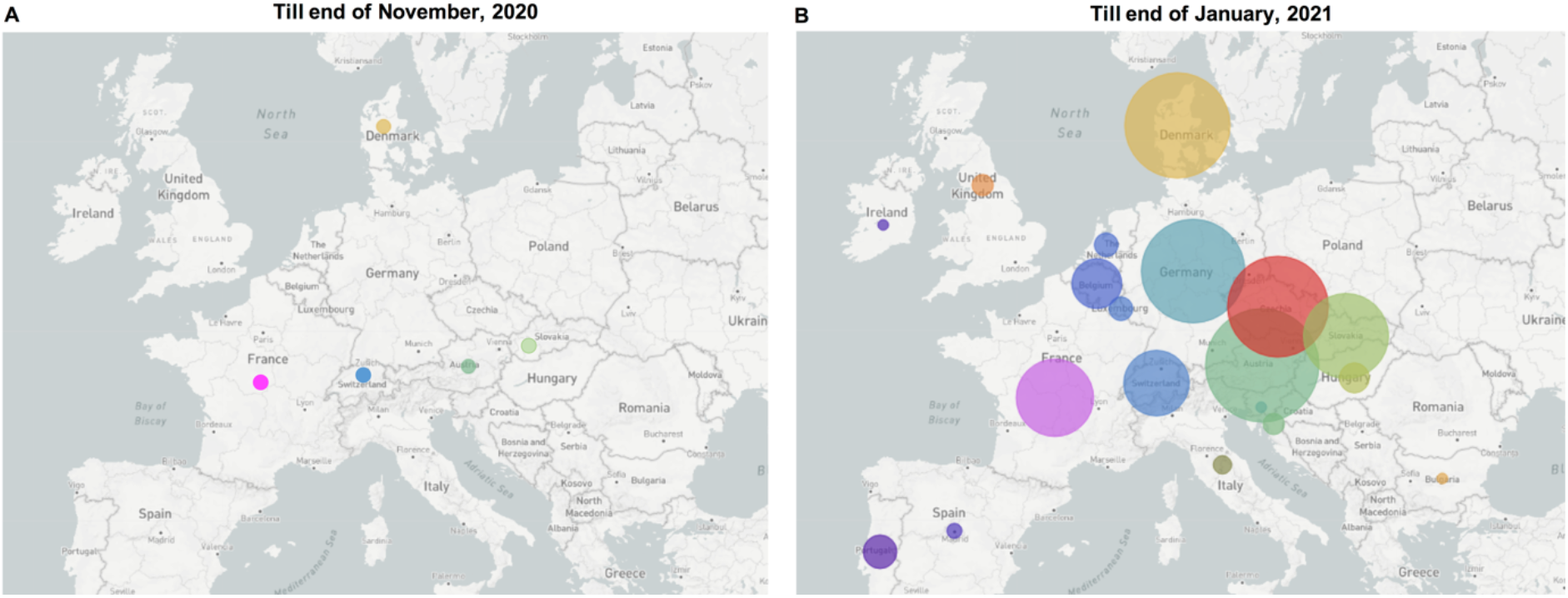
The emergence and spreading condition of this B.1.1.7 sub-lineage at the end of November 2020 (A) and at the end of January 2021 (B). Each circle represents the number of genomes reported from each country.

In the Czech Republic, the first sample of this B.1.1.7 sub-lineage was reported at the beginning of January 2021, which was also the first B.1.1.7 variant being reported locally. However, it should be pointed out that in December only 39 SARS-CoV-2 samples were sequenced, which means the early spreading of the B.1.1.7 sub-lineage might be missed out owing to the low sampling density, and it could be that in late December the B.1.1.7 sub-lineage already started community transmission. The B.1.1.7 sub-lineage in the Czech Republic was possibly from one of the Czech neighbor countries, such as Slovakia or Austria, where the B.1.1.7-N:G204P-ORF8:68stop sub-lineage was detected much earlier.

### 4. The COVID-19 pandemic waves in early 2021 in the Czech Republic were associated with the expansion of the B.1.1.7-N:G204P-ORF8:68stop sub-lineage

Since this B.1.1.7 sub-lineage was dominant in the Czech Republic and accounted for around 95% B.1.1.7 related cases, to investigate the impact of the spreading of this B.1.1.7 sub-lineage on the COVID-19 pandemic in the Czech Republic, we analyzed the details of the COVID-19 pandemic conditions between December 2020 to April 2021 in the Czech Republic and the corresponding SARS-CoV-2 lineage development during the same period. For COVID-19 pandemic, following a second wave from September 2020 till November 2020 which was known to be caused by the spreading of other previously existing lineages (20, 21), a sharp wave occurred between December 2020 to January 2021 (Fig. 4A). For SARS-CoV-2 lineage development, between December 2020 to January 2021, the prevalence of B.1.1.7 (B.1.1.7 samples/all sequenced samples) increased from 0% in late December 2020 to ∼ 60% in late January 2021, replacing the majority of other lineages (Fig. 4B) (Note: as mentioned in the results section 3, the early growth of the B.1.1.7 sub-lineage might be missed out from the genome surveillance owing to the low sampling density in December), suggesting the major driving force for the sharp wave was the quick expansion of the B.1.1.7 variant, which was shown to be more transmissible than previously existing other lineages (2-4). Although the January wave was curbed temporarily by some countermeasures, after the January peak, the 7-day incidence rate was kept at a high level (above 400), and reached another peak in early March (Fig. 4A&C) along with the further expansion of B.1.1.7. Across the time period between January to April 2021, the ratio of the B.1.1.7-N:G204P-ORF8:68stop sub-lineage to all B.1.1.7 in the Czech Republic was between 91% to 100% (Fig. 4C).

**Fig. 4.**
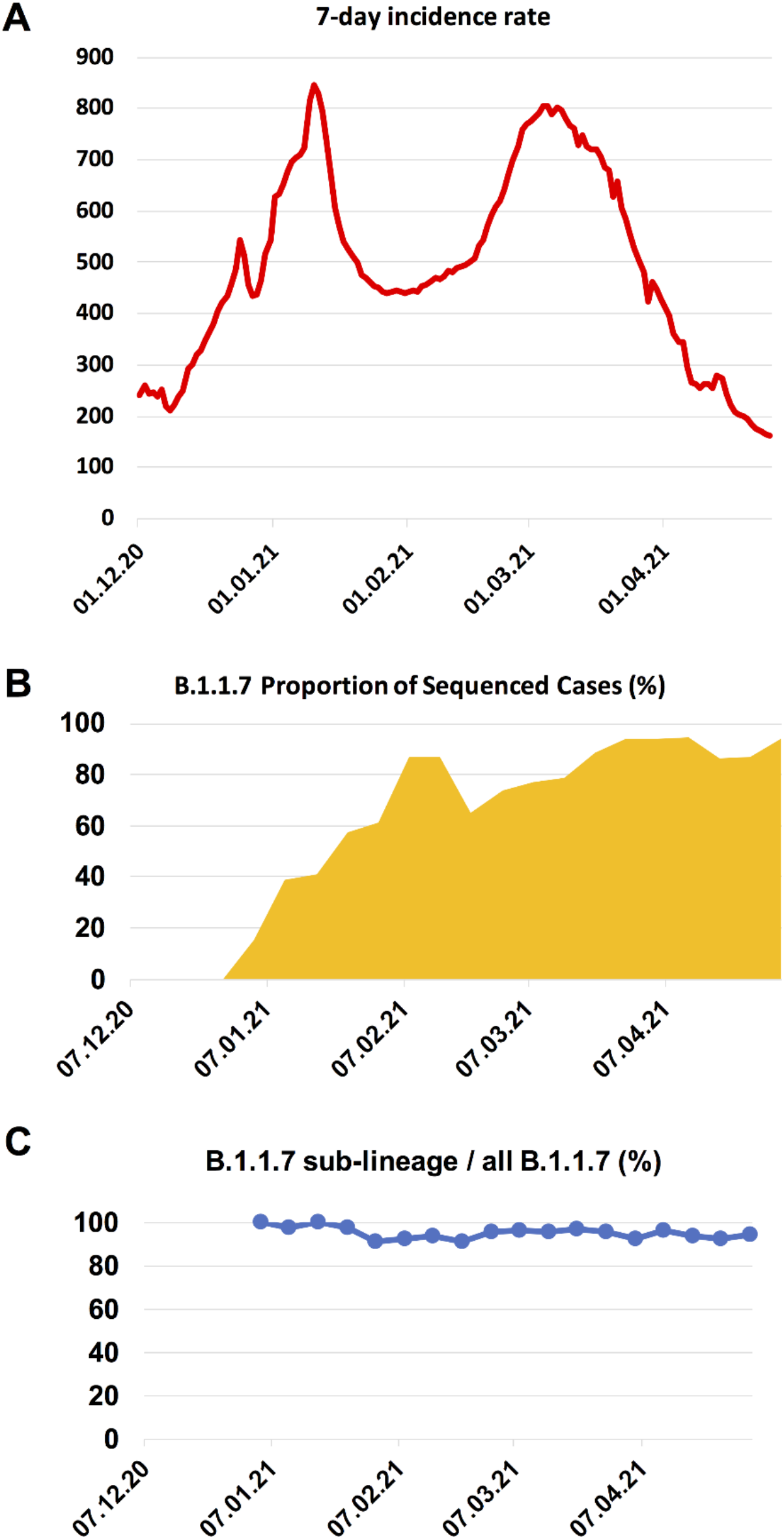
The COVID-19 pandemic waves in early 2021 and the expansion of the B.1.1.7 in the Czech Republic. **A**. 7-day incidence rate per 100K inhabitants in the Czech Republic. **B**. B.1.1.7 proportion of sequenced cases in the Czech Republic in each week, starting from the week 1^st^ – 7^th^ December, 2020. The B.1.1.7 lineage rose rapidly in early 2021, replacing the majority of other lineages (shown as the white blank space) present during this time period. **C**. The ratio of the B.1.1.7-N:G204P-ORF8:68stop sub-lineage to all B.1.1.7 in each week, starting from the first week when B.1.1.7 was detected.

The prevalence of B.1.1.7 in the Czech Republic in January was much higher than that of most other European countries. For example, the prevalence of B.1.1.7 in Poland in late January 2021 was only around 10% (Fig. S1A). Besides, in most other European countries, such as in Poland (Fig. S1B), the B.1.1.7 wave peaked later at the end of March or the beginning of April (https://qap.ecdc.europa.eu/public/extensions/COVID-19/COVID-19.html#countrycomparison-tab).

### 5. Signature mutations

#### 5.1 Signature mutations of the B.1.1.7-N:G204P-ORF8:68stop sub-lineage

Within this sub-lineage, the most common signature mutations were the same as B.1.1.7 signature mutations, with the additional **N:G204P** and **ORF8:68stop** mutations. As described below, there were other novel common spike mutations detected in a small portion of samples from this sub-lineage.

#### 5.2 Further mutation accumulation in the B.1.1.7 sub-lineage

Through extensive investigation of this B.1.1.7 sub-lineage, we detected one variant with further accumulated mutations in the spike protein on the genetic background of the B.1.1.7-N:G204P-ORF8:68stop sub-lineage, characterized by two further extra spike mutations: D138Y and A701V (Fig. 5A). This variant was mainly detected in Germany, accounting for 0.07% of all B.1.1.7 in Germany as of June 30, 2021. However, it is noteworthy that the two extra mutations carried by this B.1.1.7 variant are shared with other VOCs. The spike D138Y is one signature mutation for the Gamma variant (22), and the spike A701V is one signature mutation for the Beta variant (23). For samples sequenced locally with vaccine documentation, this variant accounted for around 50% B.1.1.7 related vaccine breakthrough cases.

**Fig. 5.**
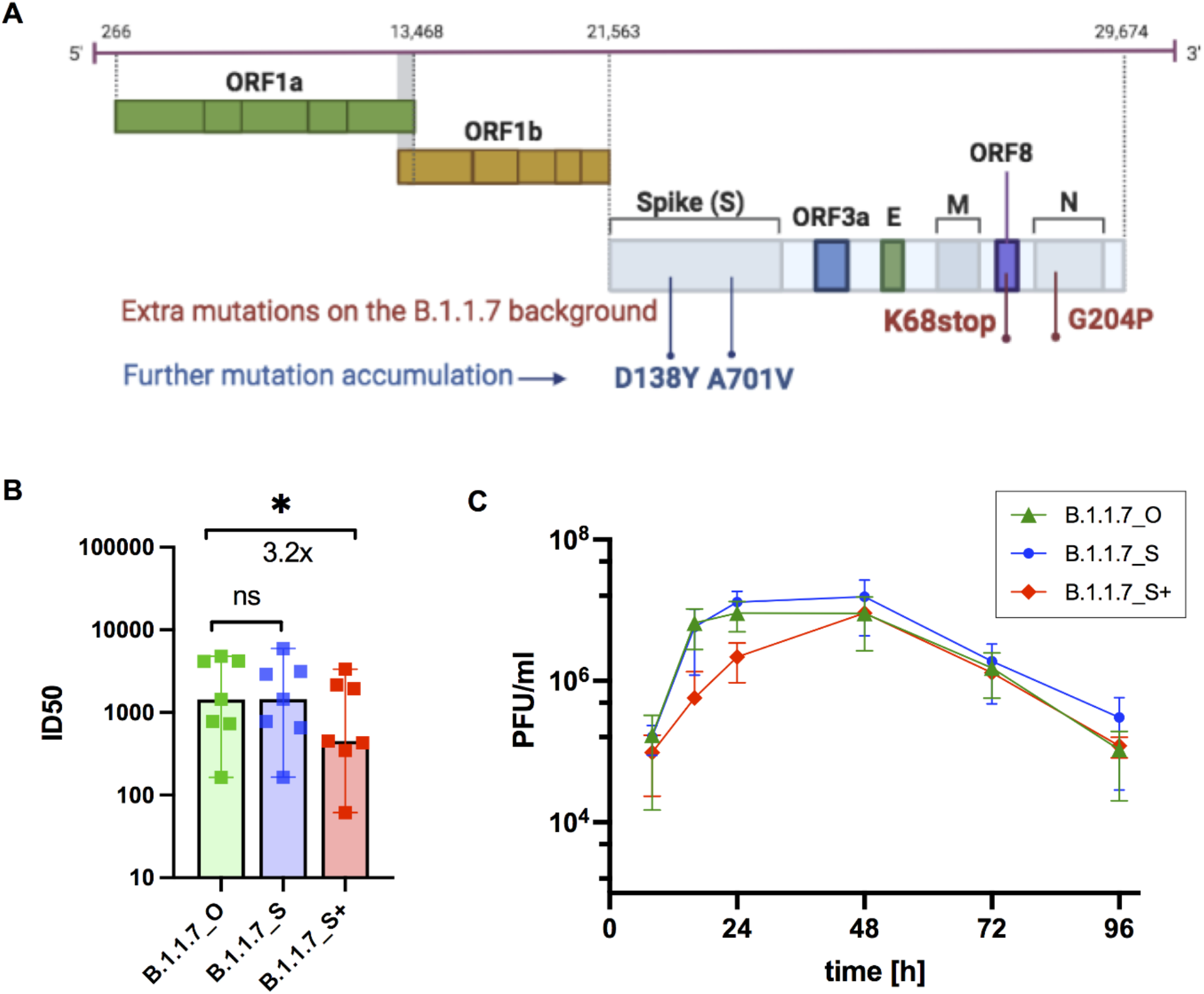
Neutralization efficacy and growth kinetics of three B.1.1.7 variants. **A**. Further mutant accumulations of D138Y and A701V in the spike protein were detected within the SARS-CoV-2 B.1.1.7-N:G204P-ORF8:68stop sub-lineage. **B&C**. Functional evaluation of virus isolates from the three B.1.1.7 variants: the originally defined B.1.1.7 (B.1.1.7_O), the B.1.1.7-N:G204P-ORF8:68stop sub-lineage (B.1.1.7_S) and the B.1.1.7-N:G204P-ORF8:68stop sub-lineage carrying two extra spike mutations D138Y and A701V (B.1.1.7_S+). **B**. Neutralization efficacy of sera from fully vaccinated individuals (n=7, BNT162b2) against active virus of the three B.1.1.7 variants. ID_50_, the serum dilution required for 50% virus inhibition. Bars represent the median ID_50_ values with 95% CI. *p<0.05 Wilcoxon matched-pairs signed rank test, ns not significant. **C**. Growth kinetics of B.1.1.7_O, B.1.1.7_S and B.1.1.7_S+ on Calu 3 cells as titrated by plaque assay. All data represent three independent experiments each with two technical replicates.

### 6. Impact of the mutations in the B.1.1.7 variants on virus propagation and antibody neutralization

We used virus isolates from the three variants B.1.1.7_O (similar to originally defined B.1.1.7), B.1.1.7_S (B.1.1.7-N:G204P-ORF8:68stop sub-lineage) and B.1.1.7_S+ (B.1.1.7-N:G204P-ORF8:68stop sub-lineage with extra spike mutations D138Y and A701V) to test their susceptibilities to vaccine elicited serum neutralizing antibodies in individuals following vaccination with two doses BNT162b2. These experiments showed a decrease of neutralization sensitivity for B.1.1.7_S+, which carries two extra spike mutations D138Y and A701V, compared to the other two B.1.1.7 variants B.1.1.7_O and B.1.1.7_S of around 3.2-fold (Fig. 5B).

To evaluate replication abilities of these three B.1.1.7 variants, we infected a lung epithelial cell line, Calu-3, with the three B.1.1.7 variant isolates. We observed a replication disadvantage for B.1.1.7_S+ compared to B.1.1.7_O and B.1.1.7_S (Fig. 5C) in the first 24 hours after infection. These data support lower replication rate and therefore lower transmissibility of B.1.1.7_S+ over B.1.1.7_O and B.1.1.7_S.

## Discussion

In this article we describe the detection, characterization, transmission, evolution and functional evaluation of the B.1.1.7-N:G204P-ORF8:68stop sub-lineage that emerged in the European continent in November 2020 and spread widely in many European countries as of 30 June 2021. This B.1.1.7-N:G204P-ORF8:68stop sub-lineage was first detected in November 2020 in Switzerland, Austria, France, Slovakia and Denmark. In late December 2020 or early January 2021, it spread to the Czech Republic and quickly became the local predominant SARS-CoV-2 variant replacing the majority of previously existing variants, and then spread to other European countries. Furthermore, we detected evolution of this B.1.1.7 sub-lineage, characterized by samples carrying two extra spike mutations A701V and D138Y on the genetic background of this B.1.1.7 sub-lineage.

With virus isolates from the three B.1.1.7 variants: the originally defined B.1.1.7 (B.1.1.7_O), the B.1.1.7-N:G204P-ORF8:68stop sub-lineage (B.1.1.7_S) and the B.1.1.7-N:G204P-ORF8:68stop sub-lineage carrying two extra spike mutations D138Y and A701V (B.1.1.7_S+), we have tested among the B.1.1.7 variants if there is any difference in replication efficiency or neutralization sensitivity against vaccine elicited serum neutralizing antibodies. The results indicated the B.1.1.7_S itself showed no replication advantage or reduced neutralization sensitivity compared to the B.1.1.7_O. This explains the co-existence of the two variants in many European countries as shown in Figure 1. However, for the B.1.1.7_S+, reduced neutralization sensitivity was observed, indicating potential for immune evasion (24-26). At the same time, the B.1.1.7_S+ isolate showed decreased replication fitness compared to the other B.1.1.7 variants, which explains its limited expansion in Europe and low ratio of detection in Germany (less than 0.1% of all B.1.1.7 as of June 30, 2021).

Furthermore, using virus genome surveillance data we have estimated the cross-country spreading pattern of B.1.1.7 in Europe in January 2021 when B.1.1.7 started being frequently detected in most European countries. We find the spreading pattern of B.1.1.7 throughout Europe was shown as high export frequencies from the major source locations, UK and Czech Republic, which is comparable to the early spreading pattern of B.1.1.7 in the UK (8) and in a few other countries at the national level (17, 18, 27).

The identification of this B.1.1.7 sub-lineage has solved an important puzzle about the spreading of B.1.1.7 in Europe. The SARS-CoV-2 lineage B.1.1.7 was first detected in the UK in late 2020, and then spread to other countries. It is commonly acknowledged that geographic distance and traffic connection to the epicenter play an important role in the spreading pattern of virus (28, 29). Usually, in the region closer to the epicenter, the virus would emerge and spread earlier. In most countries in Europe, the B.1.1.7 wave peaked at the end of March or early April. Among European countries, Czech Republic is geographically farther away from the UK than most other countries, but in January 2021 the B.1.1.7 had already become the predominant SARS-CoV-2 lineage there, which was earlier than most other countries. It was difficult to explain the contradiction between the geographical distribution and the time line of the B.1.1.7 spreading, but the spreading of this B.1.1.7 sub-lineage in the European continent since November 2020 has provided one answer for that.

Moreover, this B.1.1.7 sub-lineage might be the only B.1.1.7 variant that was not restrained by the local control measures in the Czech Republic and went on with community transmission in last December or early January 2021. Perhaps driven by various transient demographic and epidemiological factors, such as opening of ski resorts and other human mobilities related with vacation or family gathering, or local high incidence rate that may lead to escalating growth rate of SARS-CoV-2, this B.1.1.7 sub-lineage rapidly increased in the Czech Republic and then spread to a variety of other countries.

The wide spread of this B.1.1.7 sub-lineage was related to the pandemic waves in the Czech Republic in early 2021, and played a role in driving the B.1.1.7 wave in various other European countries, such as Austria, Slovakia, Germany and Denmark. These findings add to the understanding about the pandemic development in Europe and could possibly help to prevent similar scenarios in future. Also, these findings emphasize the importance of international collaboration on virus mutant surveillance, not only for SARS-CoV-2, also for other epidemic viruses or bacteria.

## Data Availability

All the SARS-CoV-2 genomes generated and presented in this study are publicly accessible through the GISAID platform (https://www.gisaid.org/). The processed SARS-CoV-2 genome data in the form of phylogenetic tree are available at https://github.com/genomesurveillance/alpha-variant-sublineage. A list of GISAID accession ID for B.1.1.7_S+ genomes from Germany is also available at https://github.com/genomesurveillance/alpha-variant-sublineage. More general information about B.1.1.7-N:G204P-ORF8:68stop sub-lineage genome number in each country during certain time period can be acquired by choosing the relevant location and collection period on the GISAID database (with searching items: VOC Alpha (variants); Submission date is before 2021-06-30; Substitutions: N_G204P and NS8_K68stop). To access sequence data from GISAID, registration with https://www.gisaid.org/ is necessary, which involves agreeing to GISAID Database Access Agreement. Biological materials (i.e. virus variant isolation) generated as a part of this study will be made available but may require execution of a materials transfer agreement.

## Acknowledgements

We thank all researchers who are working around the clock to generate and share genome data on GISAID (http://www.gisaid.org). We specifically thank colleagues at the Institute of Medical Microbiology and Virology, University Hospital Carl Gustav Carus, for their work in performing SARS-CoV-2 sample testing and sequencing sample preparing, and we thank the Dresden concept Genome Center for their sequencing efforts. We thank the Robert Koch Institute for the data management and sharing. Parts of this study were supported by a grant from the German Ministry of Health to A.D. (project LüSeMut). We thank Dr. R. Weidemann and A. Zabzinski for help with this project. We thank all the collaboration partners who contributed to the project LüSeMut.

## Data availability

All the SARS-CoV-2 genomes generated and presented in this study are publicly accessible through the GISAID platform (https://www.gisaid.org/). The processed SARS-CoV-2 genome data in the form of phylogenetic tree are available at https://github.com/genomesurveillance/alpha-variant-sublineage. A list of GISAID accession ID for B.1.1.7_S+ genomes from Germany is also available at https://github.com/genomesurveillance/alpha-variant-sublineage. More general information about B.1.1.7-N:G204P-ORF8:68stop sub-lineage genome number in each country during certain time period can be acquired by choosing the relevant location and collection period on the GISAID database (with searching items: VOC Alpha (variants); Submission date is before 2021-06-30; Substitutions: N_G204P and NS8_K68stop). To access sequence data from GISAID, registration with https://www.gisaid.org/ is necessary, which involves agreeing to GISAID’s Database Access Agreement. Biological materials (i.e. virus variant isolation) generated as a part of this study will be made available but may require execution of a materials transfer agreement.

## Code availability

Data processing and visualization was performed using publicly available software, primarily RStudio v1.3.1093. Phylogenetic maximum likelihood (ML) and time trees were constructed using the SARS-CoV-2-specific procedures taken from github.com/nextstrain/ncov.

